# Targeted Connectomic Neuromodulation of the Orbitofrontal Cortex To Treat Obsessive-Compulsive Disorder

**DOI:** 10.64898/2026.05.26.26354163

**Authors:** Emma Anderson, Audrey Kist, Zachary D Simon, Jason Raj, Sneha Ray, Dani Astudillo, Natalie Becker, Tenzin Norbu, Starlette Khim, Dennis Lambert, John Alvarez, Kelly Kadlec, Anusha B Allawala, Alexandra Tremblay-McGaw, Jessica Verhein, Caroline A Racine, Pierre Naldec, Ahmad Alhourani, Keaton Piper, Joline M Fan, Doris D Wang, Ankit N Khambhati, Kristin K Sellers, Phillip A Starr, Leo P Sugrue, Edward F Chang, Andrew D Krystal, A Moses Lee

## Abstract

Pathological activity within frontal cortical circuits is common in many neuropsychiatric disorders, such as obsessive-compulsive disorder (OCD). We developed an invasive brain mapping protocol in which temporary electrodes are implanted in candidate sites to identify personalized stimulation targets that can acutely relieve OCD symptoms. We found that stimulation within segments of the anterior limb of the internal capsule (ALIC) focally suppressed the structurally and functionally connected region of prefrontal and cingulate cortex. By leveraging the topographic organization of the ALIC, we reversibly inactivated frontal cortical sites with ALIC stimulation to determine which cortical regions are necessary for sustaining OCD symptoms. Stimulation of ventral capsule (VC) near the globus pallidus within the ALIC was associated with suppression of lateral orbitofrontal cortex activity and acute and long-term improvements in OCD symptoms. These results provide a paradigm for leveraging ALIC topography to deliver targeted connectomic neuromodulation to frontal cortex to treat neuropsychiatric disorders.

## Introduction

Pathological activity within prefrontal cortex (PFC) and anterior cingulate cortex (ACC) is common in many neuropsychiatric disorders. For example, contemporary circuit models of obsessive-compulsive disorder (OCD) hypothesize that persistent hyperactivity across a recurrent cortico-striato-thalamo-cortical (CSTC) circuit encompassing the orbitofrontal and cingulate cortex, striatum, and associated thalamic regions give rise to patterns of recurrent intrusive, distressing thoughts and repetitive behaviors [1, 2]. Yet, our ability to reliably and reversibly target neuromodulation to these frontal cortical circuits remains limited, creating a methodological barrier to developing new treatments for these conditions.

For OCD, deep brain stimulation (DBS) has emerged as a treatment for severe and refractory cases [3]. A number of evidence-based DBS targets exist for OCD such as the anterior limb of the internal capsule (ALIC) adjacent to the nucleus accumbens (NAc) or bed nucleus of the stria terminalis (BNST)[4-6]. The ALIC consists of a heavily myelinated fiber bundles containing descending and ascending frontal cortical fibers to the thalamus and brainstem[7-9]. These ALIC fibers are embedded in the NAc as small myelinated fascicles and run adjacent to the BNST. The ALIC is known to have a topographic organization[10]. Fibers from ventral prefrontal cortical regions travel more ventrally in the capsule compared to those connected to more dorsal prefrontal or cingulate cortical regions. In this way, ALIC DBS has the potential to target neuromodulation to a wide range of prefrontal circuits from a central topographically organized white matter hub. Elucidating general principles regarding how the prefrontal cortex (PFC) and anterior cingulate cortex (ACC) respond to ALIC stimulation could have important therapeutic implications not only for treating OCD, but also for treating other neuropsychiatric disorders in which these cortical circuits are dysregulated.

However, DBS outcomes for OCD remain variable with an approximately 60% response rate across DBS targets [6, 11, 12]. Attempts to optimize DBS for treating OCD have been limited by the lack of understanding regarding its underlying circuit-based mechanism of action, which could be used to improve DBS targeting and identify biomarkers of circuit-based target engagement. For this reason, we developed an invasive brain mapping protocol that aimed to identify personalized neuromodulation targets to optimize therapeutic outcomes in severe, refractory OCD. We initially developed this approach to identify antidepressant stimulation responses, adapting methods commonly used in epilepsy to inform surgical treatment of epilepsy[13-15]. In our participants with refractory OCD, we implanted temporary intracranial electrodes across the CSTC circuit in a set of evidence-based candidate targets of neuromodulation for OCD[1, 2]. We performed stimulation mapping to identify therapeutic sites that acutely reduced OCD symptoms while avoiding sites that led to adverse effects. The invasive brain monitoring stage provided us with the rare opportunity to characterize the acute effect of stimulation on the CSTC circuit, using a multi-modal approach combining intracranial recordings and pre-operative imaging. We then implanted chronic DBS electrodes in the top personalized therapeutic sites to implement a novel combined multi-site neuromodulation protocol with the goal of achieving a more rapid and consistent relief in OCD symptoms across subjects than has previously been possible.

For this study, we aimed to address several questions with the invasive brain mapping phase of our study: 1) Can stimulation of topographically-organized ALIC subregions modulate specific frontal circuits? 2) What are the specific components of frontal cortical circuitry within the CSTC that need to be engaged with DBS to relieve OCD symptoms? 3) Where should stimulation be targeted in the ALIC to elicit acute and long-term benefit in OCD symptoms? Here, we present data from the first four participants of our multi-stage FDA-approved (IDE G240038) clinical trial (ClinicalTrials.gov NCT06347978) using invasive brain mapping to guide personalized, multi-lead DBS to treat OCD.

## Results

### Stimulation Mapping Identifies Acutely Therapeutic VC Sites Across Participants

In four participants, stereoencephalography electrodes were implanted in the orbitofrontal cortex (OFC), cingulate cortex, NAc, BNST, and subthalamic nucleus (STN). We conducted extensive safety testing and evoked potential stimulation mapping across the CSTC network. We then conducted short 5-minute stimulation trials across CSTC targets to assess safety and preliminary efficacy using visual analogue scales (VAS) of Obsession, Compulsions, and OCD-specific distress as well as clinical observations while intracranial recordings were collected. VAS of Depression, Anxiety, and Energy were also collected. The most effective stimulation sites that were not associated with adverse effects were evaluated in randomized sham-controlled testing using 20-minute stimulation blocks, with VAS surveys administered every 5 minutes (Fig. 1B). In each of our participants, we found that the VAS-OCD composite score (calculated as the sum of the VAS obsessions, compulsions, and OCD-specific distress) correlated with the Y-BOCS I and II during period of time before and after the invasive brain mapping stay (Supplementary Fig. 1). To quantify the effects of stimulation, we calculated the magnitude of change in the mean VAS-OCD composite score relative to sham (Fig. 1C). We also calculated the mean change in VAS for the obsession, compulsion, and OCD-related distress domains as well as for depression, anxiety, and energy across participants for stimulation trials across distinct sites (Supplementary Fig 2).

**Figure 1.**
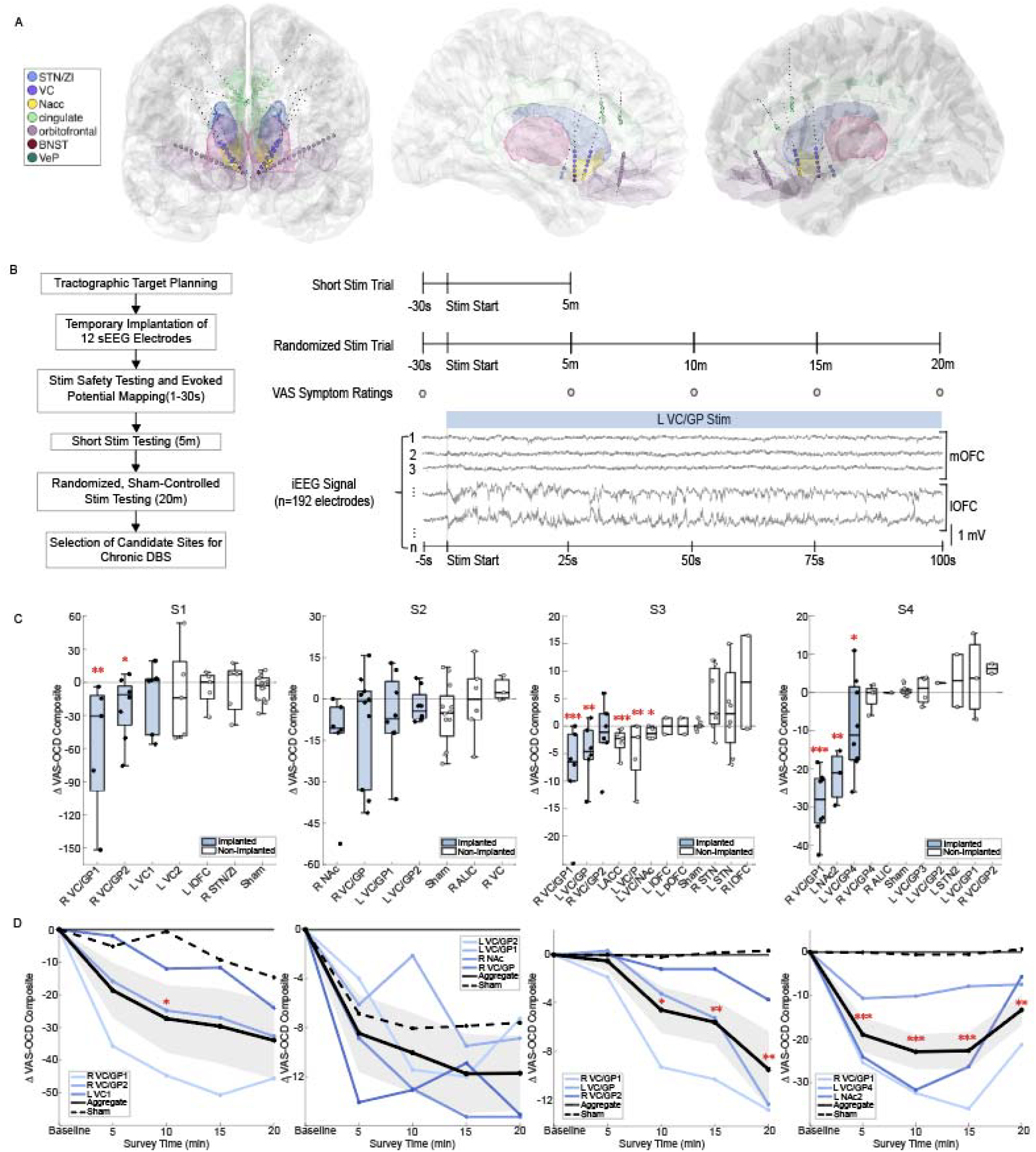
Invasive Brain Mapping Identifies Ventral Capsule as an Acutely Therapeutic Stimulation Target for OCD. **(A)** Representative Localization of sEEG leads in Study Participants and Stimulation Mapping Paradigm. Contacts located in regions of interest for OCD are shown in color; other contacts are shown in black. Top: frontal view; middle row: left hemisphere; bottom row: right hemisphere. **(B)** Diagram of study paradigm and stimulation paradigm. **(C)** Change in OCD-composite score during stimulation of different targets and sham across the four participants. Stars indicate a significant change from sham control (permutation test, *n*= 10,000, * p < 0.05). **(D)** Time course of OCD-composite score relative to stimulation onset for acutely therapeutic implanted sites across all four participants.

Across subjects, the largest and most consistent improvements in the VAS-OCD composite score could be observed with stimulation of sites within the ventral capsule (VC) often located adjacent to the globus pallidus. (Fig. 1C). Most sites associated with significant improvements in OCD symptoms were not associated with significant changes in VAS-Depression, Anxiety, or Energy, nor were they associated with significant adverse effects (Supplementary Fig. 3). These ventral capsule/globus pallidus (VC/GP) targets were subsequently implanted in each of the participants for chronic DBS.

We next sought to characterize the time course of the VAS-OCD composite response for each of the acutely therapeutic VC targets and sham stimulation. Across all participants and therapeutic VC targets, the average improvement in VAS-OCD composite score with stimulation increased with longer durations of stimulation (Fig. 1D).

### Leveraging the Topographic Connectivity of the ALIC to Suppress Activity within Distinct Regions of Frontal Cortex

We next characterized the structural and functional connectivity of our VC/GP targets relative to other ALIC sites in the NAc and dorsal ALIC (dALIC). Prior studies have demonstrated that white matter tracts connected to regions of the prefrontal and cingulate cortex are topographically organized within the ALIC. We performed seed-based diffusion tractography from three stimulation sites in the ALIC: 1) NAc, 2) VC/GP, and 3) dorsal ALIC (dALIC). We found that: 1) the NAc was structurally connected to the medial OFC (mOFC), 2) the VC/GP was structurally connected to the lateral OFC (lOFC) and anterior cingulate cortex (aCin), and 3) dALIC was structurally connected to the dorsolateral prefrontal cortex (dlPFC), including middle frontal gyrus (MFG) and superior frontal gyrus (SFG), as well as the dorsal cingulate cortex (dCin) (Fig 2A). Across participants, the acutely therapeutic VC/GP stimulation sites that were implanted for chronic DBS had dense connectivity to both lOFC, aCin, dorsomedial prefrontal cortex, medial thalamus, and brainstem (Supplementary Fig 4 and 5).

**Figure 2.**
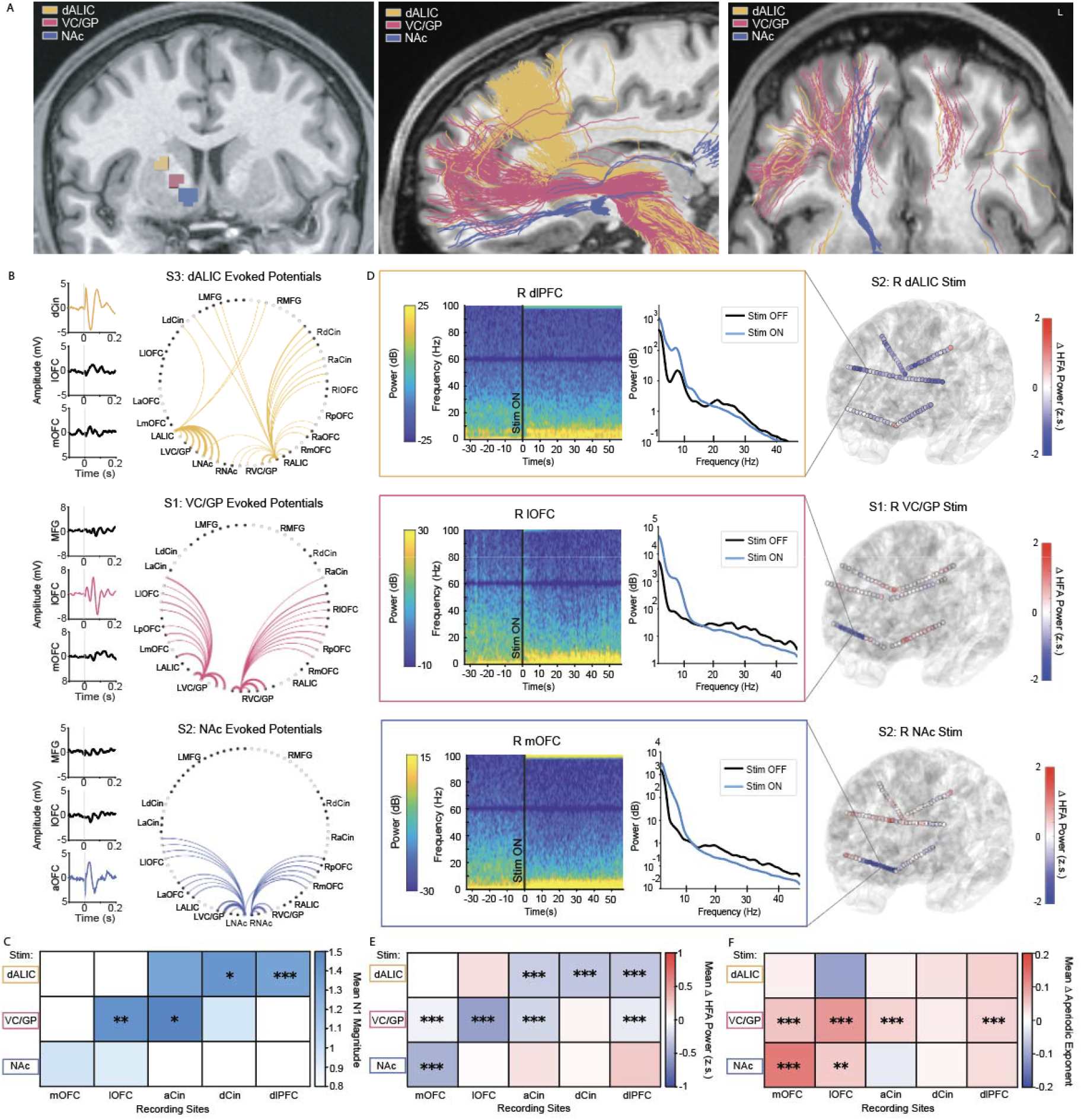
Topographically Organized Structural and Directed Function Connectivity Predict the Region of Prefrontal Cortex Suppressed by ALIC Stimulation. **(A)** Example tractography from ROIs in the NAc (bottom), VC/GP (middle), and dALIC (top). **(B)** Examples of EP waveforms elicited in prefrontal regions from single-pulse stimulation of NAc, VC/GP, and dALIC (left). Example EP connectivity across prefrontal cortex with single-pulse stimulation of NAc, VC/GP, and dALIC (right). **(C)** Average EP response in prefrontal regions with single-pulse stimulation in NAc, VC/GP, and dALIC. **(D)** Example spectrograms and power spectra from prefrontal recording sites in response to 100Hz trains of stimulation within the NAc, VC/GP, and dALIC (left). Examples of cortical HFA suppression in response to 100Hz trains of stimulation within the NAc, VC/GP, and dALIC (right). **(E)** Average HFA change in prefrontal regions in response to 100Hz stimulation in NAc, VC/GP, and dALIC. **(F)** Average aperiodic change in prefrontal regions in response to 100Hz stimulation in NAc, VC/GP, and dALIC. Stars indicate a significant change with stimulation relative to sham (permutation tests, *n*= 10,000, * p < 0.05, ** p < 0.01, *** p < 0.001).

We investigated whether functional connectivity assessed using single-pulse evoked potentials (SP-EP) from the 1) NAc, 2) VC/GP, and 3) dALIC recapitulated the topographic organization of white matter connectivity observed with tractography. We focused our analysis on the N1 component (10-50 ms) of the SP-EP, representing more direct connections between the stimulation target and the cortical recording site. Similar to the tractography, we observed in representative EP recording sessions that: 1) NAc SP-EP selectively elicited responses in mOFC, 2) VC/GP SP-EP selectively elicited responses in lOFC and aCin, and 3) dALIC SP-EP elicited response in aCin, dCin, and dlPFC (Fig 2B). We observed this topographic organization across all the participants with: 1) average N1 SP-EP magnitudes from NAc being greater for mOFC compared to other sites, 2) average N1 SP-EP magnitudes from VC/GP being significantly greater for lOFC and aCin compared to other sites, and 3) average N1 SP-EP magnitudes from dALIC being significantly greater for dlPFC compared to other sites (Fig 2C). Across all participants, the acutely therapeutic VC/GP stimulation sites that were implanted for chronic DBS had dense connectivity to lateral OFC (Supplementary Fig 6).

We next examined the electrophysiological consequences at cortical recording sites in response to 5 and 20-minute trains of stimulation at the NAc, VC/GP, and dALIC sites using intracranial recordings (Fig. 2D). Our analysis focused on cortical effects where the impact of ALIC stimulation artifacts was less apparent compared to subcortical recording sites closer to the site of stimulation. In particular, we measured the high frequency activity (HFA; 30-90 Hz), an index of local neuronal population firing and aggregate synaptic activity, and the aperiodic exponent, reflecting the broadband 1/f slope linked to excitation/inhibition balance, across all cortical recording sites[16]. In representative cortical recordings, stimulation induced a focal cortical suppression of HFA accompanied by a shift from high to lower-frequency dynamics measured as a larger aperiodic exponent across ALIC stimulation sites in all four participants (Fig 2D). These spectral changes were visually apparent as a ‘focal slowing’ of local field potential activity within PFC and ACC (Fig 1B, Supplementary Fig 7) and persisted even after the end of the stimulation train (Supplementary Fig 8).

The spatial locus of the suppression of HFA was dependent on the stimulation sites within the ALIC and resembled the structural/EP connectivity of the stimulation target (Fig 2E). Of all three ALIC sites, NAc stimulation elicited the strongest suppression of HFA in mOFC, which was statistically significant relative to sham. VC/GP stimulation elicited the strongest suppression of lOFC, which was statistically significant relative to sham. dALIC stimulation elicited the strongest suppression of aCin, dCin, and dlPFC, which were statistically significant relative to sham (Fig 2E).

Likewise, the anatomical location of the increase in the aperiodic exponent was also dependent on the stimulation sites within the ALIC and resembled the structural/EP connectivity of the stimulation target (Fig 2F). Of all three ALIC sites, NAc stimulation elicited the largest enhancement in aperiodic exponent in mOFC, which was statistically significant relative to sham. VC/GP stimulation elicited the strongest increase in aperiodic exponent in lOFC, which were statistically significant relative to sham. dALIC stimulation elicited the largest increase in aperiodic exponent within the dCin. Together, these findings demonstrate that stimulation within discrete sites within the ALIC enables anatomically specific modulation of prefrontal and cingulate regions.

### Orbitofrontal Cortex Activity Specifically Tracks OCD Symptom Improvement with Stimulation

We next sought to identify electrophysiological signatures of acute OCD symptom improvement with stimulation. We focused on subcortical stimulation sites in the NAc, BNST, VC, ALIC, and STN that most strongly modulated symptoms and investigated whether electrophysiological activity in the frontal cortex correlated with acute therapeutic responses (Fig 3A). We again examined changes in HFA and aperiodic exponent with stimulation in subcortical targets. We then correlated these electrophysiological effects with changes in the VAS-OCD composite score across stimulation targets during 20-minute randomized sham-controlled trials.

**Figure 3.**
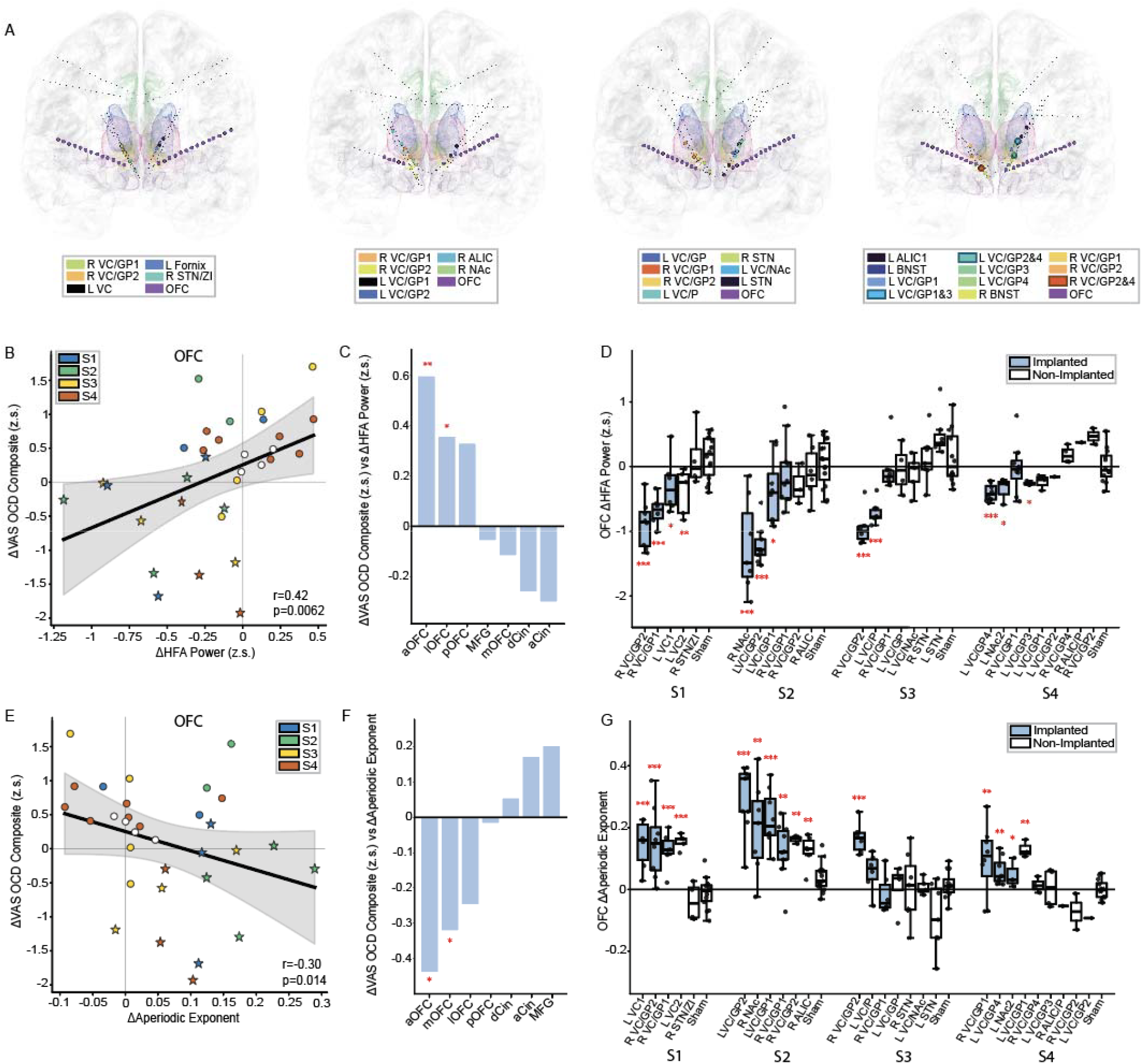
OFC Suppression Selectively Tracks Acute Improvement in OCD Severity. **(A)** Location of subcortical stimulation and OFC recording sites across all four participants. **(B)** Correlation between change in HFA power in OFC and change in OCD composite score. Stars indicate acutely therapeutic implanted targets. **(C)** Correlation between changes in HFA and VAS-OCD composite scores across prefrontal regions. **(D)** Change in OFC HFA across subcortical stimulation targets. **(E)** Correlation between change in the aperiodic component of the power spectra in OFC and change in OCD composite score. Stars indicate acutely therapeutic implanted targets. **(F)** Correlation between changes in HFA and VAS-OCD composite scores across prefrontal regions. **(G)** Change in the aperiodic components of the OFC power spectra across subcortical stimulation targets.

We found that suppressions in HFA within the OFC ipsilateral to the stimulation site were significantly correlated with reductions in the VAS-OCD composite score (r = 0.42, p = 0.0062, permutation test) (Fig 3B). The correlation between OFC HFA suppression and symptom change was stronger than the corresponding correlation at other cortical recording sites (Fig 3C). More specifically changes in the anterior and lateral OFC HFA significantly correlated with changes in the VAS obsessions, compulsions, and OCD-related distress symptom subdomains in addition to VAS-Depression (Supplementary Fig 9). Changes in aOFC HFA were also significantly correlated with VAS-Depression and Anxiety while changes lOFC HFA were significantly correlated with depression (Supplementary Fig 9). We next examined these effects at the level of individual stimulation targets. We observed significant suppression of ipsilateral OFC HFA with stimulation at eleven of the twelve VC targets that were eventually implanted (permutation test, active vs sham stimulation, p < 0.05) (Fig. 3D).

We also observed that shifts in the ipsilateral OFC power spectra towards a steeper aperiodic exponent were significantly correlated with reductions in the VAS-OCD composite score (r=−0.30, p = 0.014, permutation test) and VAS-Obsessions (Fig 3E, F, Supplementary Fig 10). Other cortical recording sites were not significantly correlated with VAS-OCD composite score. In addition, a shift in the OFC power spectra towards a steeper aperiodic exponent was observed at ten of the twelve VC targets that were eventually implanted (permutation test, active vs sham stimulation, p < 0.05) (Fig 3G).

### Clinical outcome after chronic DBS

In the second stage of our protocol, three participants were implanted with a four lead DBS system with bilateral implantable pulse generators targeted to the optimal therapeutic DBS targets and regions where chronic sensing may identify a biomarker of OCD symptoms. In the first three subjects, who underwent DBS surgery, the top three VC sites associated with acute therapeutic responses during the invasive brain mapping phase were implanted for stimulation along with a cortical sensing site in either orbitofrontal or cingulate cortex (Fig. 4A, Supplementary Fig. 11). The fourth participant is currently awaiting DBS implantation. All three subjects experienced a rapid reduction of OCD symptoms meeting responder criteria (>35% improvement) at their next appointment one or two weeks following initial DBS programming of all three VC sites based upon reduction in the Y-BOCSII (S1: 38%, S2: 41%, S3 : 49%) and Y-BOCSI (S1: 38%, S2: 42%, S3 : 39%) compared to baseline. These benefits were sustained across all three participants up until the most recent follow up appointment (Fig 4B).

**Figure 4.**
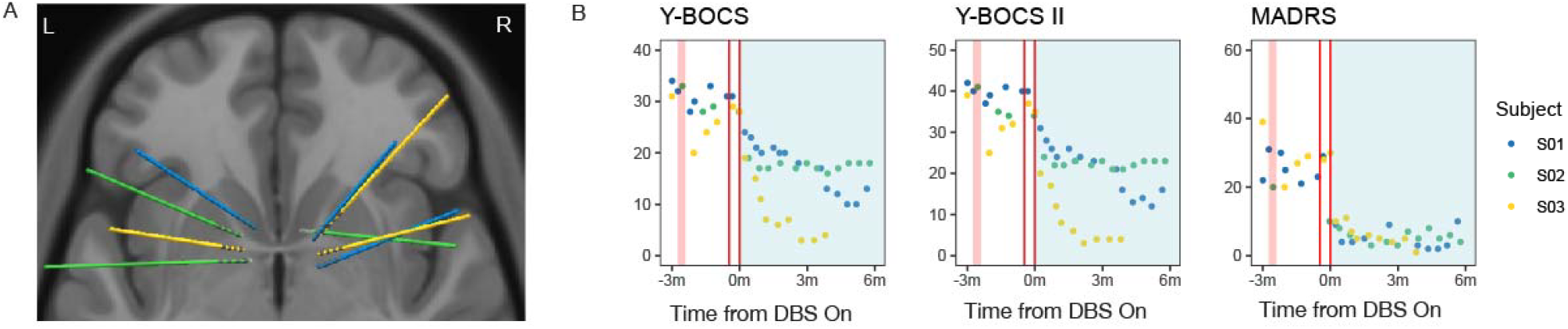
DBS Lead Placement and Outcomes from Personalized DBS Targeting. **(A)** Lead placement for Subject 1 (blue), 2 (green), and 3 (yellow). **(B)** Long-term clinical outcomes from personalized DBS across subjects. Y-BOCS-I (left), Y-BOCS-II (middle), and MADRS (right) aligned to onset of initial programming. Invasive brain mapping phase (shaded red region) took place 2 months prior to DBS surgery (first red vertical line). Initiation of DBS programming (second vertical red line, corresponding to t = 0) took place 1-2 weeks after DBS implant.

## Discussion

Here, we describe the stimulation results from our invasive brain mapping paradigm in four clinical trial participants with severe, refractory OCD. By mapping the connectivity along the ALIC, we identified that the ALIC contains a topographic functional map of the frontal cortex that can be identified using diffusion tractography and short-latency SP-EP responses[8, 10]. Importantly, stimulation within the ALIC strongly suppressed activity within the structurally and functionally connected regions of frontal cortex, as assessed by a decreases in HFA or increase in focal aperiodic slowing.

Across all participants, the stimulation sites associated with the largest acute improvements in OCD symptom were located within the lateral VC located adjacent to the globus pallidus and superior to traditional DBS targets within the NAc or BNST. These VC/GP sites rapidly reduced OCD symptoms within a period of 20 minutes and were in most cases not associated with significant changes in mood symptoms in contrast to the traditional NAc or BNST targets. By monitoring the effect that subcortical stimulation had on cortical electrophysiological activity, we observed that suppression of OFC activity specifically tracked improvement in OCD symptoms. In turn, VC/GP sites occupied a ‘sweet spot’ within the ALIC with dense tractographic and EP connectivity to the ipsilateral OFC, and stimulation of VC/GP sites strongly suppressed OFC activity. Finally, long-term chronic DBS at these VC/GP targets led to rapid and sustained clinical improvement in OCD symptoms.

In this paradigm, the HFA suppression and increases in the aperiodic exponent of the power spectra serve as electrophysiological measures of suppression and normalization of pathological cortical activity with ALIC stimulation. This ‘focal slowing’ of cortex with stimulation in many ways mimics the electrographic signature of a localized lesion[17]. These results are consistent with the model that ALIC DBS acts as a reversible ‘information lesion’[18]. The ability to reversibly inactivate specific regions of PFC and cingulate cortex on demand using targeted stimulation within the topographically organized ALIC has therapeutic implications for other neuropsychiatric disorders [8]. Our results suggest that ALIC connectomic mapping combined with these electrophysiological measures of target engagement could be used to develop a general closed-loop paradigm for neuromodulation to take offline disorder-specific cortical circuitry [19-22]. Prior studies have targeted the ALIC region for the treatment of refractory depression [13, 23, 24] and chronic pain [22, 25]. It is possible that different portions of the ALIC may need to be targeted to treat OCD, mood disorders, or chronic pain conditions. For example, the portion of the ALIC adjacent to the NAc, which is structurally connected to the subgenual cingulate cortex, mOFC, and amygdala, has been associated with improvement in mood across multiple studies [8, 23, 26, 27]. In contrast, central ALIC, projecting to ACC and centromedian thalamus, may be a better target for suppressing pain-related circuitry [28]. In future work, it should be possible to determine if targeting sub-components of the ALIC can serve as a general approach to treating a variety of neuropsychiatric disorders.

These results are also consistent with a model in which OCD symptoms are mediated by excessive activity within the OFC and its associated CSTC circuit[1]. In this model, therapeutic VC/GP stimulation blocks white matter transmission to suppress input into the OFC from thalamus and brainstem, resulting in a spectral shift from high to low frequencies and an associated improvement in clinical OCD symptoms. In this way, HFA suppression and increases in the aperiodic exponent of the power spectra may reflect a normalization of pathological excessive OFC activity via DBS. These findings are also consistent with prior fMRI studies from our group using a DBS cycling paradigm where we demonstrated that therapeutic DBS suppressed OFC activity as well as other components of the CSTC circuit, and literature demonstrating that capsulotomies are an effective treatment for OCD [29, 30].

Several recent connectomic studies have highlighted the importance of specific white matter tracts as predictors of long-term symptom improvement in OCD. The structural connectivity from the therapeutic VC/GP sites that we have identified across our four participants largely resembles the predictive white matter tracts identified in these prior studies[2, 31-34]. These tracts are components of the ALIC consisting of the OFC connections to the medial thalamus through the anterior thalamic radiation. Our findings provide an electrophysiological account for how connectomic targeting in this region can be leveraged to suppress and normalize excessive OFC activity associated with OCD symptoms [19]. By jamming axonal communication through a white matter structural hub, focal VC/GP stimulation can effectively exert broad inhibitory effects across OFC.

Our results bear some similarities to the results of Nho, et al. in which acutely therapeutic stimulation of ventral basal ganglia sites was associated with a decrease in OCD-related distress and anteromedial OFC low gamma, although, with some key differences[35]. First, Nho, et al. identified the nucleus accumbens-ventral pallidum (NAc-VP) grey matter complex as an optimal site of therapeutic stimulation whereas our therapeutic target is anatomically superior to this portion of the ALIC. In general, our target was associated with a relatively specific improvement in OCD symptoms, which merged slowly over 20-minutes of testing and only modest effects on mood (Supplementary Fig 2). In contrast, more ventral components of the ALIC, including the neighboring NAc and VP are often associated with substantial mood improvements that can be observed more quickly [13]. These regions are also structurally connected to distinct components of the prefrontal cortex with the NAc-VP being more connected with the anteromedial OFC whereas our VC/GP site was connected with lateral OFC and anterior cingulate cortex. Likewise, Nho, et al observed suppression of narrow-band low gamma in the anteromedial OFC with NAc/VP stimulation whereas we observed broadband suppressions in HFA in the more lateral aspects of the OFC with VC/GP stimulation. Lastly, we observed relatively rapid and large responses with chronic stimulation of our VC target with chronic DBS. The clinical outcomes of chronic NAc-VP stimulation remain to be seen.

The current study has several limitations. First, these findings were identified in a cohort of four participants. While the connectivity with and suppression of OFC with therapeutic VC stimulation were consistent features across all subjects, data from additional subjects will be required to determine if it is a robust predictor of DBS therapeutic response. Second, our investigations largely focused on the effect of VC stimulation at cortical sites such as OFC due to the presence of stimulation artifacts at many subcortical recording sites near the stimulation sites. This does not rule out an important role for subcortical structures in mediating therapeutic effects. Lastly, it remains to be determined if the biomarkers of therapeutic response observed during the invasive brain monitoring will generalize to the naturalistic context of the home environment. This is a question we aim to address in subsequent phases of the study using longitudinal intracranial recordings acquired with our DBS sensing device[36, 37].

Based upon these findings, we propose that a personalized clinico-electro-radiographic approach for targeting DBS to the VC could be used to optimize outcomes for treating OCD. This multi-modal approach would utilize personalized diffusion tractography to identify VC implantation sites based on tracts within the ALIC projecting to the OFC. During implantation, a second cortical sensing lead could be placed in the OFC for the purpose of verifying suppression of orbitofrontal activity during awake stimulation testing to aid in identifying therapeutic stimulation along with monitoring OCD symptoms [38]. Our results suggest that electrophysiological markers of OFC suppression such as HFA suppression and aperiodic exponent increases were less variable across stimulation trials than the improvements in the VAS, providing a potentially more reliable prediction of therapeutic effects than self-report or clinical assessments can provide on their own. Such a testing procedure could be performed during a sub-acute hospital stay similar to the present design of our clinical trial[39] or potentially even intra-operatively, thereby eliminating the need for inpatient stimulus response-mapping which would dramatically reduce clinical burden and cost, thus allowing for wider adoption of personalized DBS for OCD in the future. Moreover, the identification of OFC suppression as a biomarker of therapeutic response paves the way for closed-loop DBS strategies as well as other non-invasive forms of neuromodulation that could lead to better outcomes with fewer off-target effects [21, 23, 40, 41].

## Data Availability

All data produced in the present study are available upon reasonable request to the authors

## Contributions

AML, ADK, and EFC initiated the work and supervised the study. KKS and PAS contributed to the conceptualization and evaluation of the study protocol. AML, AK, KKS, EA, ZDS, JA, KK, JMF, ABA, TN, DA, NB, ATM, JV collected the data. EA, AK, ZS, ANK, JR, and SR analyzed the data. EA, AML, and AK drafted the manuscript. ADK and EFC reviewed the initial draft of the manuscript. AML recruited and clinically managed the participants. EFC, PAS, AA, and KP performed the surgeries. All authors revised the manuscript, approved the work, and take responsibility for its integrity.

## Acknowledgements

This work was supported by the Foundation for OCD Research (A.M.L.), One Mind (A.M.L.), National Institutes of Mental Health (NIMH) K23MH125018 (A.M.L.), NIMH R21MH130914 (A.M.L), Foundation for the NIH (A.M.L), and NARSAD Young Investigator grant from the Brain & Behavior Research Foundation (A.M.L.).

