## Supplemental Methods and Figures for "Targeted Connectomic Neuromodulation of the Orbitofrontal Cortex To Treat Obsessive-Compulsive Disorder"

**Supplementals:**

### Methods

#### Clinical Trial, Ethics Approval and Consent

All participants provided written informed consent prior to enrollment in this SEEG-guided personalized DBS clinical trial for OCD (ClinicalTrials.gov identifier: NCT06347978), which was approved by the Institutional Review Board at UCSF and by the U.S. Food and Drug Administration. The study was conducted in accordance with the principles of the Declaration of Helsinki. Participants additionally provided written authorization for the use of personal health information for research purposes and for the publication of study-related images.

**The clinical trial consists of three stages. Stage 1 involves invasive brain mapping and stimulation using sEEG to characterize neural activity within brain regions implicated in OCD symptomatology. Stage 2 consists of the surgical implantation of up to four permanent DBS leads at locations personalized based on the Stage 1 investigation, followed by programming and optimization of stimulation parameters to reduce symptoms. Stage 3 comprises a double-blind, randomized crossover phase, followed by an open-label follow-up period.**

#### Electrode Implantation

Twelve-16 contact sEEG electrodes (PMT Corporation or AdTech) were implanted bilaterally targeting: VC/NAc, VC/BNST, anterior medial subthalamic nucleus/zona incerta (amSTN/ZI), OFC, ACC, and DCC. Subcortical leads used 2.5mm center-to-center contact spacing and cortical leads used 3.5mm spacing for subject 1, while later subjects were implanted with 3mm center-to-center spaced leads for all targets. Surgical targeting combined anatomic landmarks and diffusion tractography in Brainlab iPlan. Post-operative CT was registered to pre-operative MRI to verify contact locations, confirmed by neuroradiologist review. Recording and stimulation procedures occurred over 10 days before electrode explantation.

#### sEEG recordings and stimulation

We performed a 3-phase extensive stimulation mapping testing during a 10-day inpatient monitoring stay. In Phase 1, we conducted safety testing with brief stimulation trains across anatomical regions of interest at currents ranging from 1-6 mA and durations of 1-300 seconds to rule out sites associated with adverse effects, such as involuntary movements, autonomic effects, excessive mood elevation concerning for mania, or spiking activity on sEEG monitoring concerning for progression to a seizure. Symptoms were assessed using visual analogue scales (VAS) for obsessions, compulsions, OCD-related distress, depression, anxiety, and energy. Top candidate sites were tested using a 20-minute randomized, sham-controlled stimulation paradigm. Both the participant and clinicians were blinded to stimulation site, with repeated trials of stimulation. A provocation paradigm was used during testing to elicit OCD symptoms if the participant was asymptomatic.

#### High-Frequency Activity (HFA) and Aperiodic Component Analyses

HFA and aperiodic spectral features were analyzed from continuous sEEG recorded during stimulation blocks (5-minute and 20-minute duration, 100 Hz, bipolar, 100 µs pulse width, 1.0 to 6.0 mA). Data were downsampled to 512 Hz, notch filtered at 60 Hz and harmonics, and bipolar re-referenced. HFA (30 to 90 Hz) was extracted via bandpass filtering, with power spectra computed at 5 second resolution. HFA power was z-scored to a 30 second pre stimulation baseline and averaged across the frequency band. Change in HFA was quantified by comparing average power 30 seconds pre stimulation to 5 minutes post stimulation onset. Power spectra from the same contacts were decomposed into periodic and aperiodic components using the Fitting Oscillations and One Over F (FOOOF) algorithm[14]. Spectra from 1 to 90 Hz were fit using a fixed aperiodic mode with peak width limits of 0.5 to 30 Hz and a minimum peak height of 0.01. Aperiodic change was quantified comparing average aperiodic exponent 30 seconds pre stimulation to 5 minutes post stimulation onset. Statistical significance was assessed via permutation testing (n = 10,000), comparing therapeutic and sham recordings.

#### DTI Connectivity

Pre-surgical 3T MRI included T1-weighted BRAVO (1mm isotropic) anatomical and HARDI diffusion imaging (b=2000s/mm², 83 directions). Diffusion data underwent noise reduction, distortion correction (TOPUP/Eddy), and multi-tissue constrained spherical deconvolution (MRtrix3, Dhollander algorithm). Stimulation contact coordinates were estimated from post-operative T1 MRI and CT co-registration (LeGUI, Rolston lab)[15]and were transformed to diffusion space via rigid registration. Spherical 2mm-radius ROIs were created at each electrode coordinate using the MarsBar toolbox in SPM. Probabilistic tractography (iFOD2, 50,000 streamlines) was filtered to only those streamlines passing through combined ROIs composed of the ROIs for each electrode in a given therapeutic configuration. Structural connectivity to therapeutic contacts was estimated from the filtered streamline sets to support electrophysiological findings.

#### Evoked Potentials Connectivity

Single-pulse electrical stimulation (bipolar, 3mA, 1Hz, 20 trials per site) was delivered to adjacent contact pairs across all regions. sEEG was recorded at 10kHz using a 256-channel Nihon Kohden system. EP responses were quantified by z-scoring voltage against 50ms pre-stimulus baseline, then measuring root-mean-square power of trial-averaged waveforms in the N1 window (10-50ms post-stimulus). EP magnitudes exceeding threshold (z>1) defined functional connectivity between stimulation sites and recording contacts.

#### Clinical Assessment

#### OCD symptoms were assessed using the Yale–Brown Obsessive–Compulsive Scale (Y-BOCS) and the Yale–Brown Obsessive–Compulsive Scale, Second Edition (Y-BOCS-II) (Goodman et al., 1989; Storch et al., 2010). Depressive symptoms were assessed using the Montgomery–Åsberg Depression Rating Scale (MADRS) (Montgomery & Asberg, 1979).

#### Because the Y-BOCS and Y-BOCS-II assess symptom severity over the one to two weeks preceding evaluation, they are not suitable for capturing rapid, within-session symptom fluctuations, such as those induced by acute stimulation during Stage 1 (invasive sEEG mapping). To assess acute symptom changes, we therefore developed visual analog scales (VAS) ranging from 0 to 100. OCD-related VAS included ratings of obsessions, compulsions, and OCD-related distress. Non–OCD-specific VAS included anxiety, depression, and energy. All VAS ratings were self-reported by participants using a tablet interface.

#### Chronic, multi-site DBS Therapy

SEEG electrodes were explanted at the end of the invasive monitoring stay. Eight weeks later, the participant underwent four-lead DBS surgery with bilateral Medtronic Percept RC implantable pulse generators at sites associated with acute therapeutic improvement. Two weeks following implantation, an open-label DBS programming phase was initiated in which DBS parameters were adjusted to achieve optimal clinical improvement.

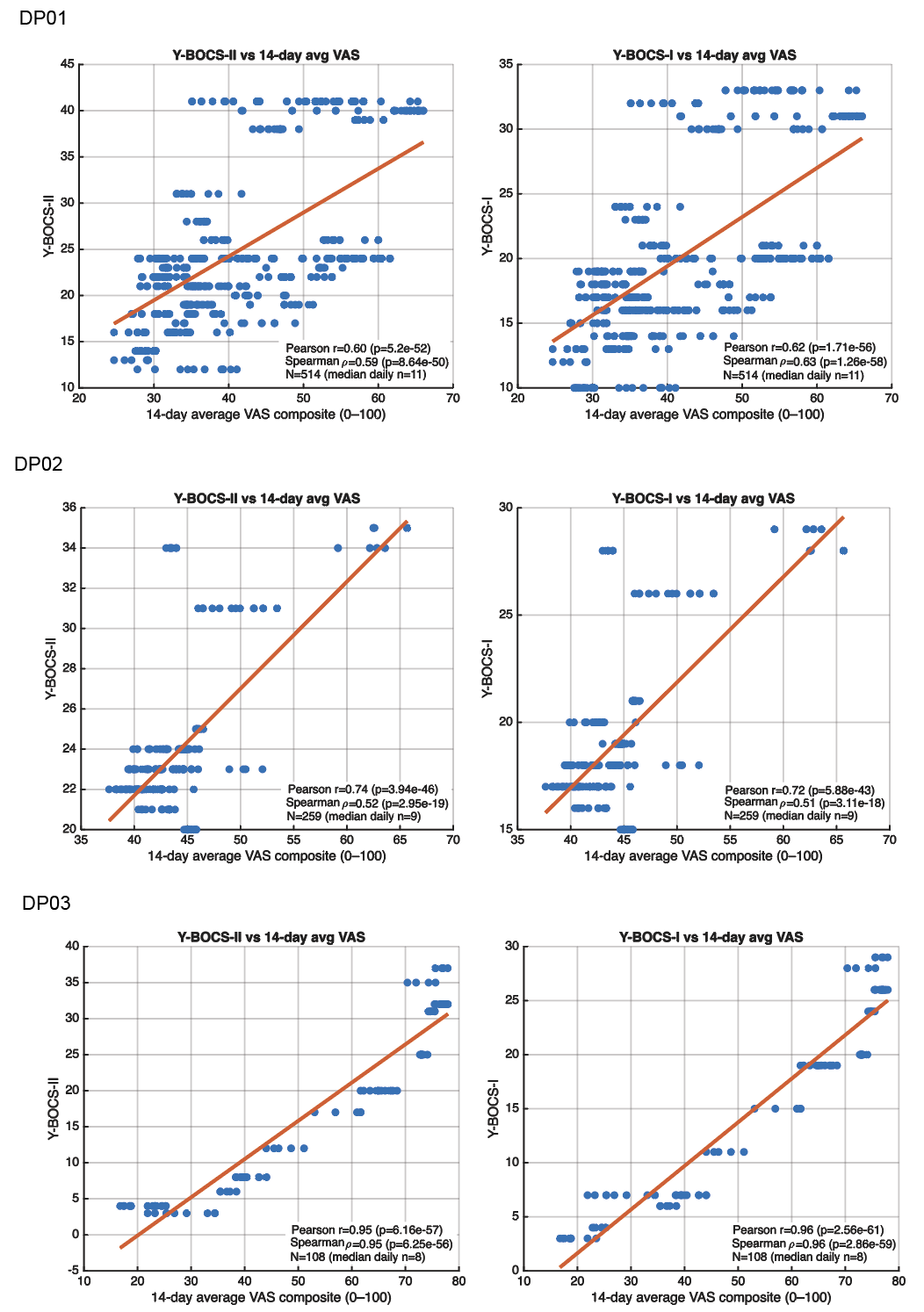

**Supp Fig 1: Validation of VAS OCD-Composite against Y-BOCS 1 and 2**. Correlation between the 14-day average VAS OCD-composite score and Y-BOCS I and II scores across subjects 1-4

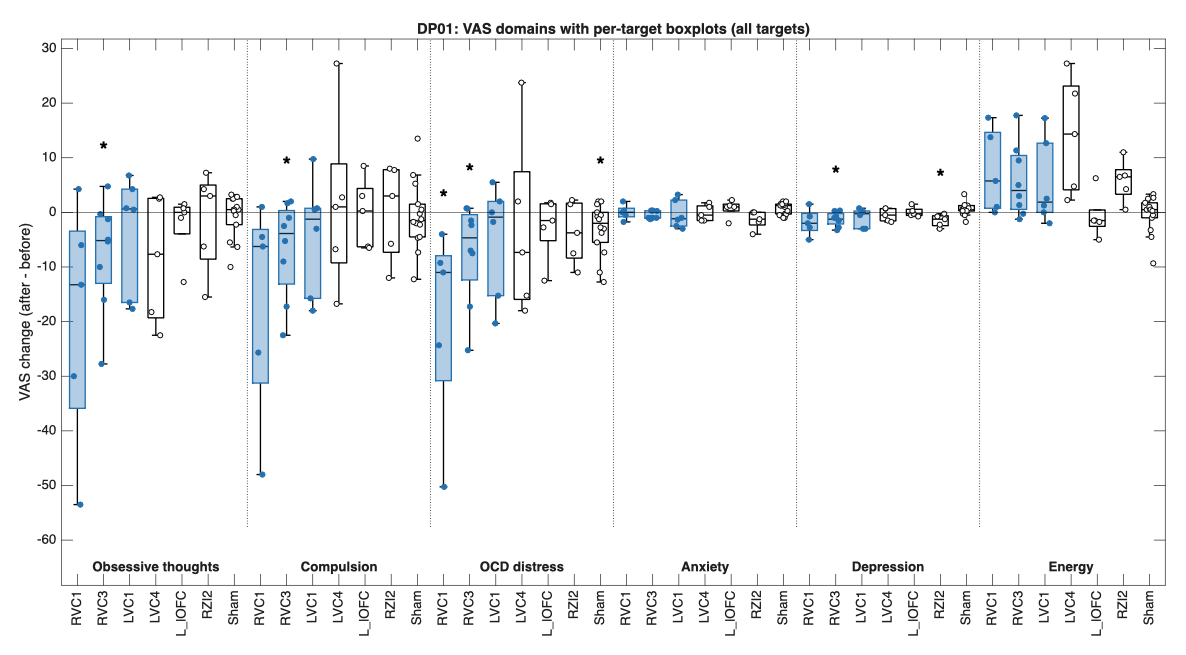

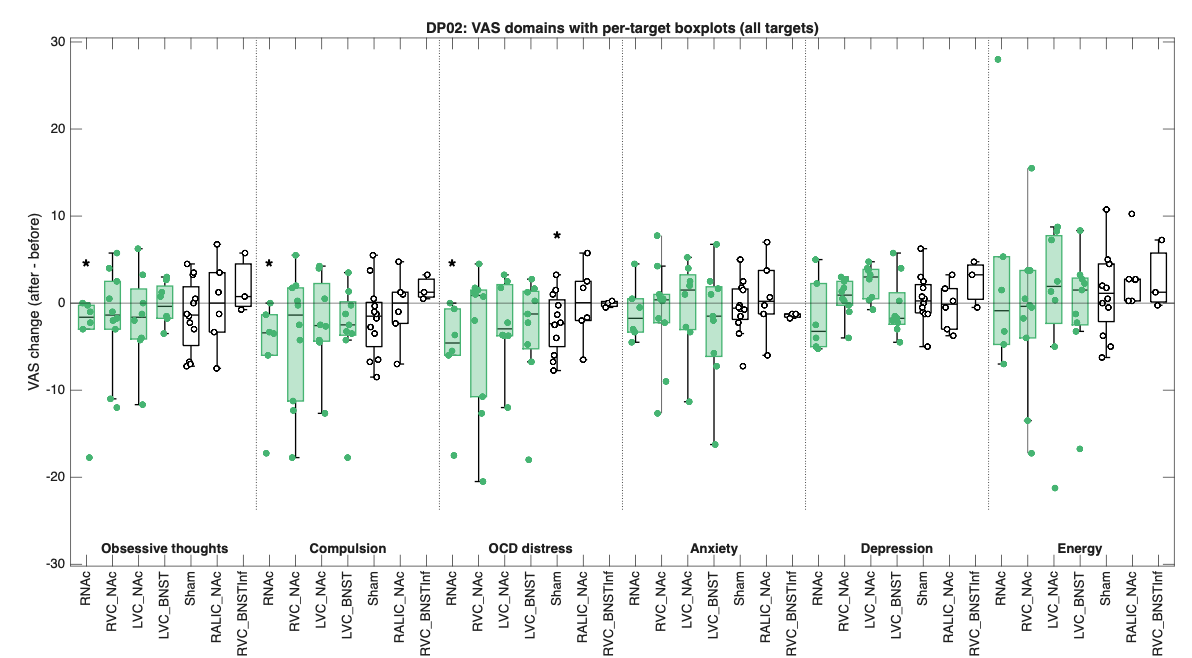

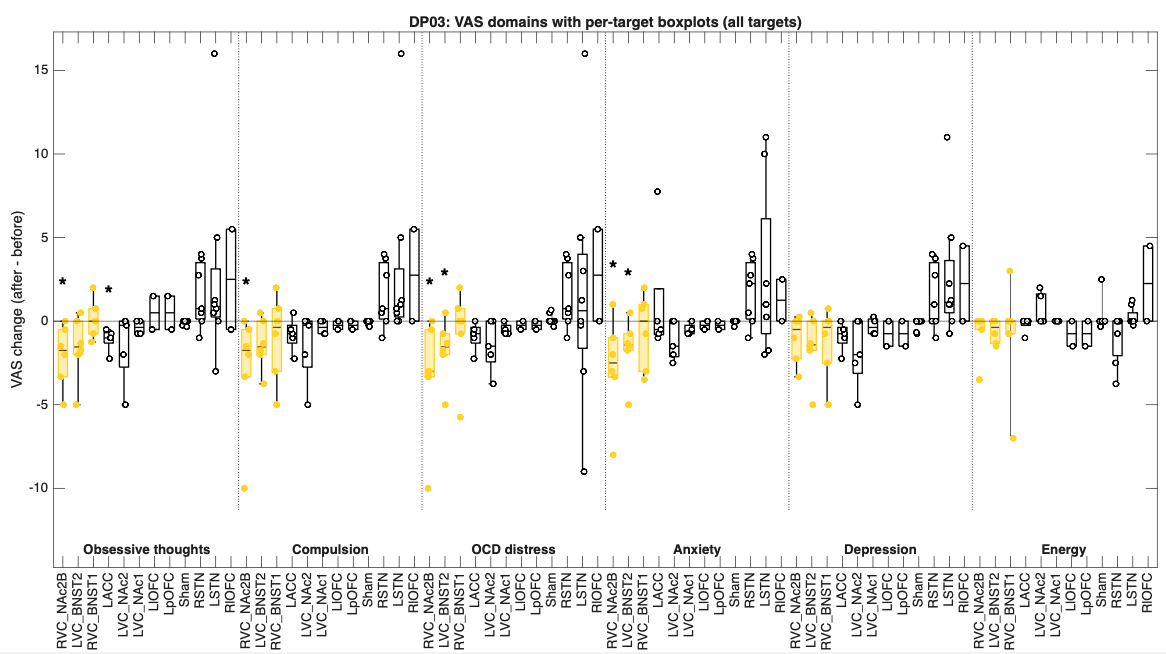

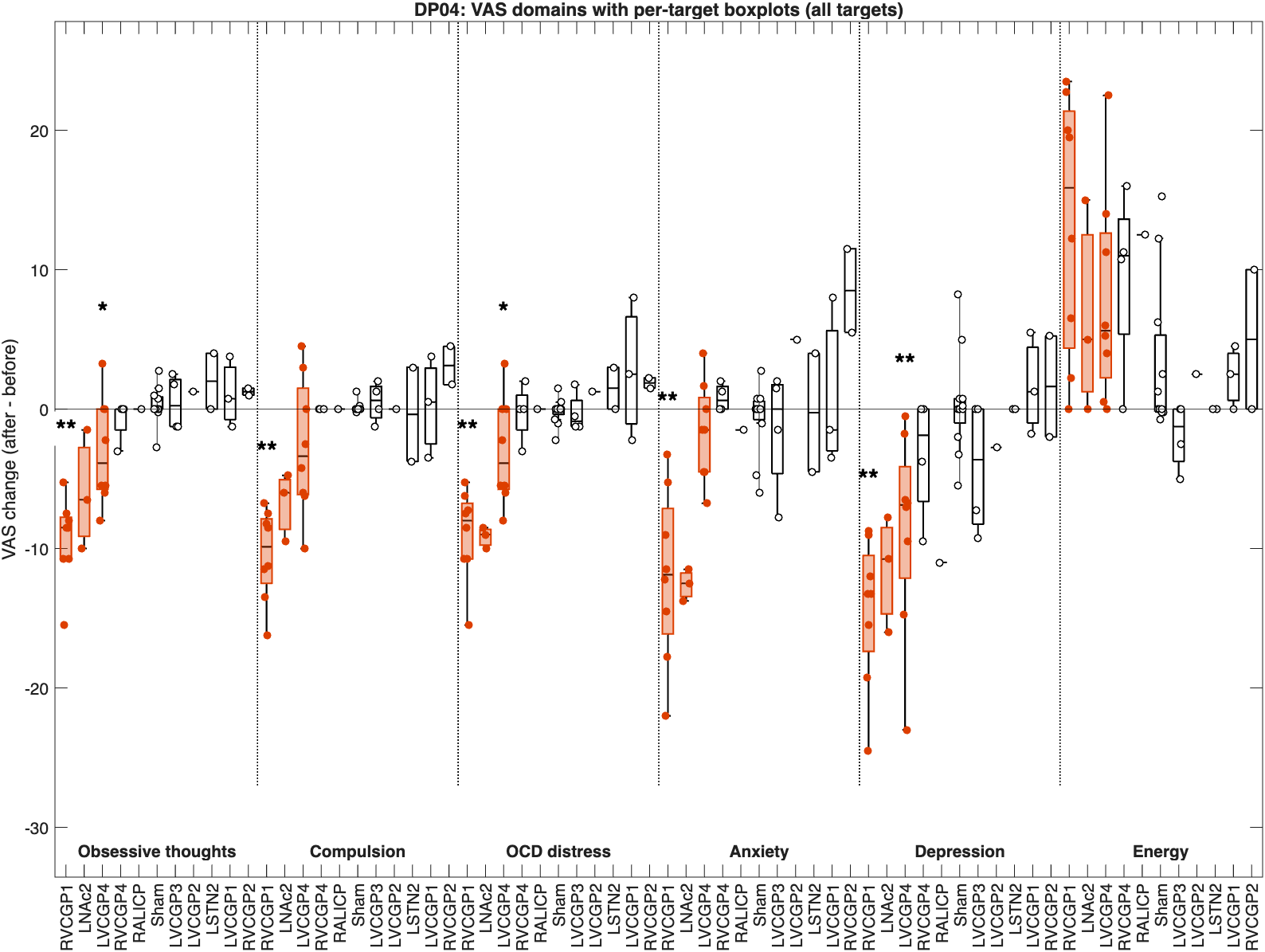

**Supp Fig 2: Changes in Symptoms with Stimulation Target Across Participants.**  Change in Visual Analog Scale (VAS) for obsessive thoughts, compulsions, OCD distress, anxiety, depression, and energy are shown for each stimulation target across subjects 1-4. Values represent the change in symptom severity relative to the baseline survey, calculated as the average score during the 20-minute stimulation block minus baseline.

| **DP01**  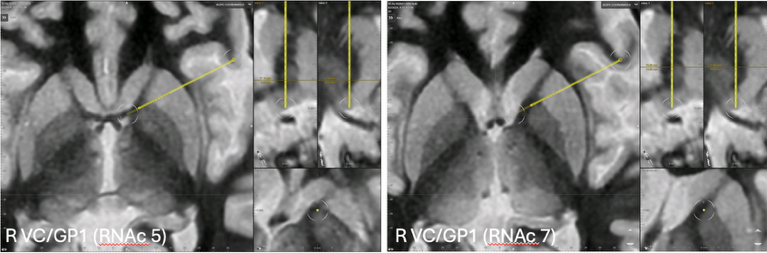  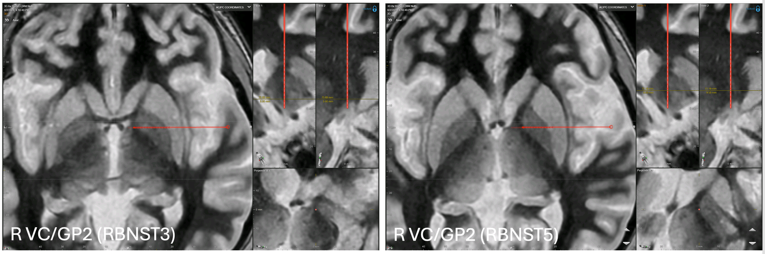  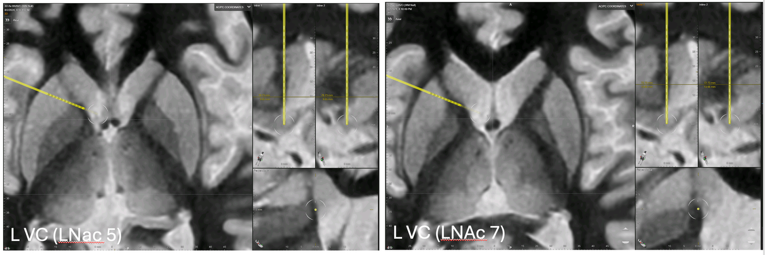 | **DP02**  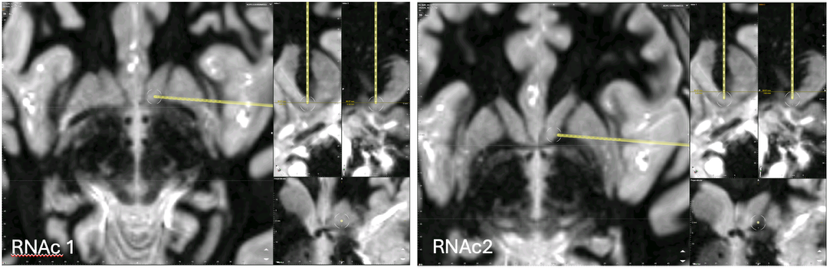  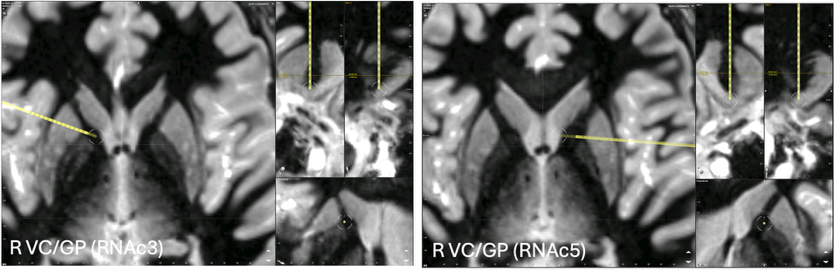  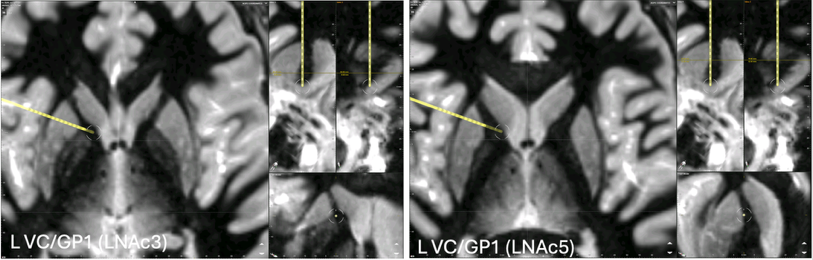  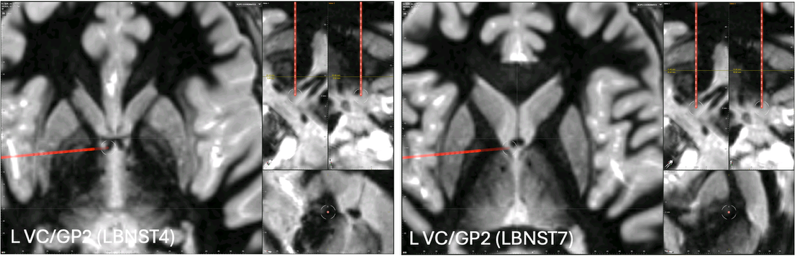 |
| --- | --- |
| **DP03**  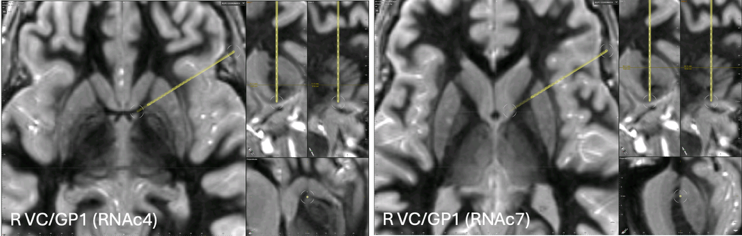  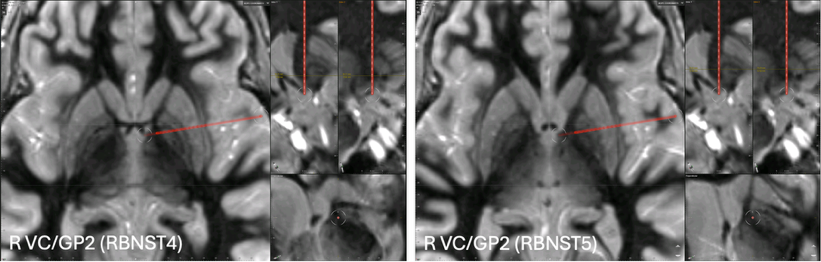  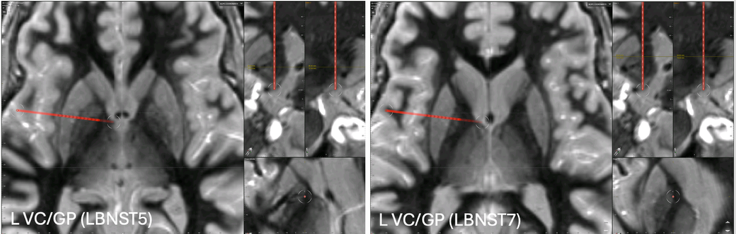 |  |

**Supp Fig 3: Location of Acutely Therapeutic Contacts across Subjects.** Horizontal and inline views showing the anatomical locations of acutely therapeutic stimulation contacts featured as bipolar pairs identified across subjects 1-4.

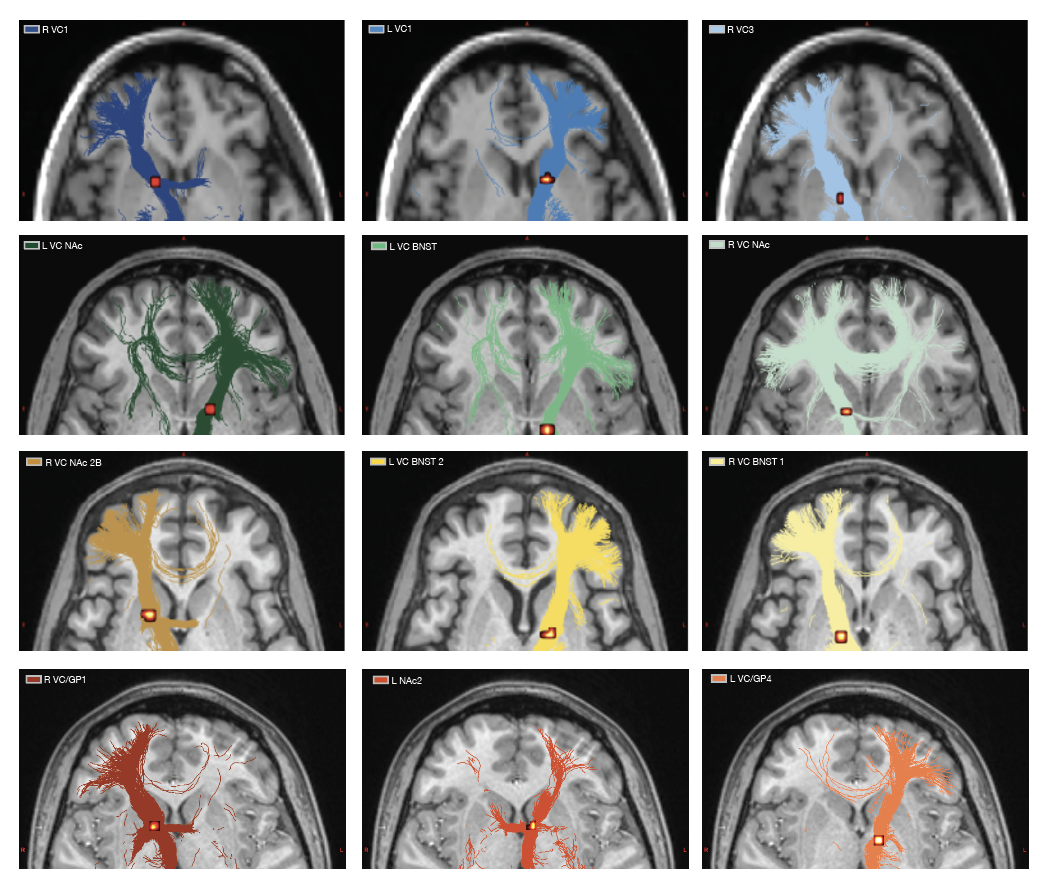

**Supp Fig 4: Horizontal View of Tractography from Acutely Therapeutic VC Stimulation Sites.** Horizontal views of diffusion tractography seeded from acutely therapeutic ventral capsule (VC) stimulation sites in subjects 1-4. Streamlines are shown for therapeutic stimulation configurations in each subject and overlaid on structural MRI. Colors correspond to individual subjects: subject 1 (blue), subject 2 (green), subject 3 (yellow), and subject 4 (red). Therapeutic contact locations were estimated from post-operative imaging and used as seed regions for probabilistic tractography (50,000 streamlines).

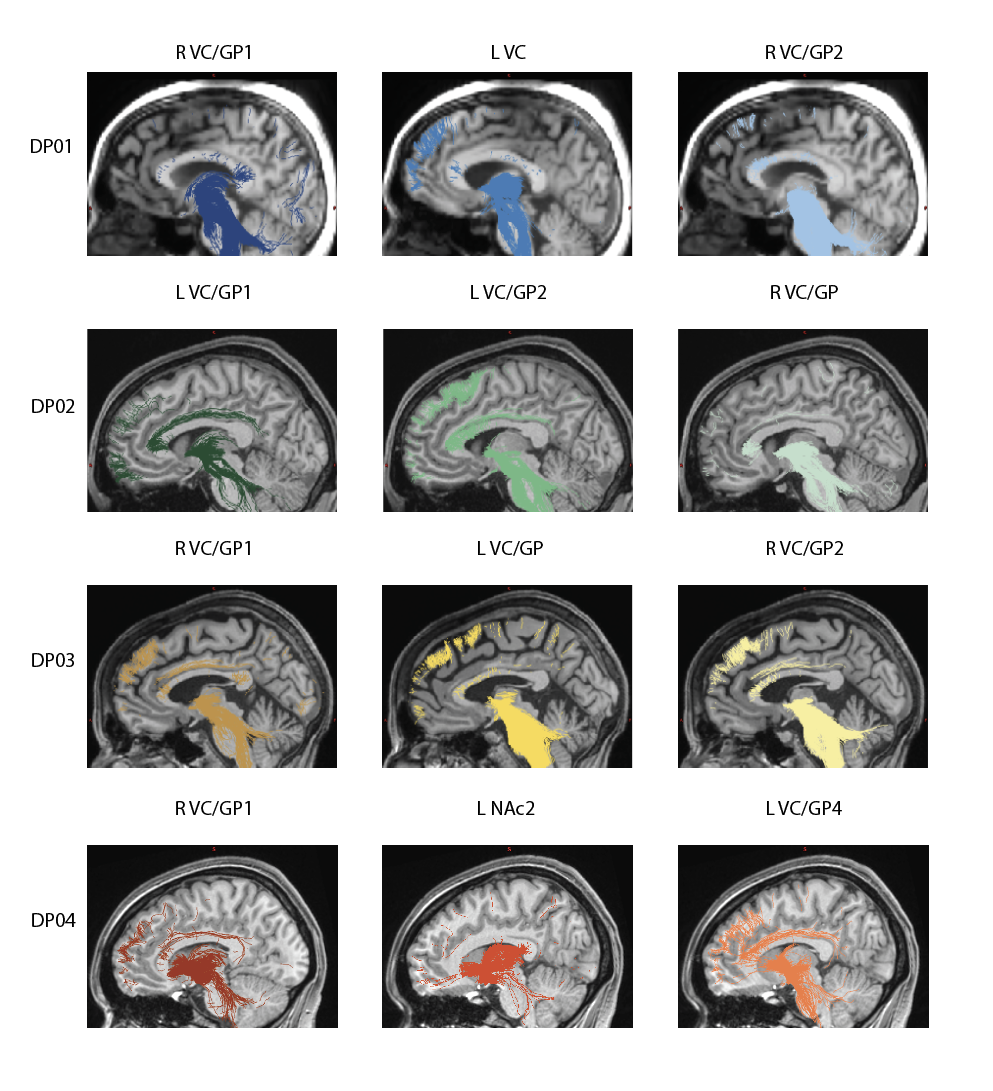

**Supp Fig 5: Sagittal View of Tractography from Acutely Therapeutic VC Stimulation Sites.** Sagittal views of diffusion tractography seeded from acutely therapeutic ventral capsule (VC) stimulation sites in subjects 1-4. Streamlines are shown for therapeutic stimulation configurations in each subject and overlaid on structural MRI. Colors correspond to individual subjects: subject 1 (blue), subject 2 (green), subject 3 (yellow), and subject 4 (red). Therapeutic contact locations were estimated from post-operative imaging and used as seed regions for probabilistic tractography (50,000 streamlines).

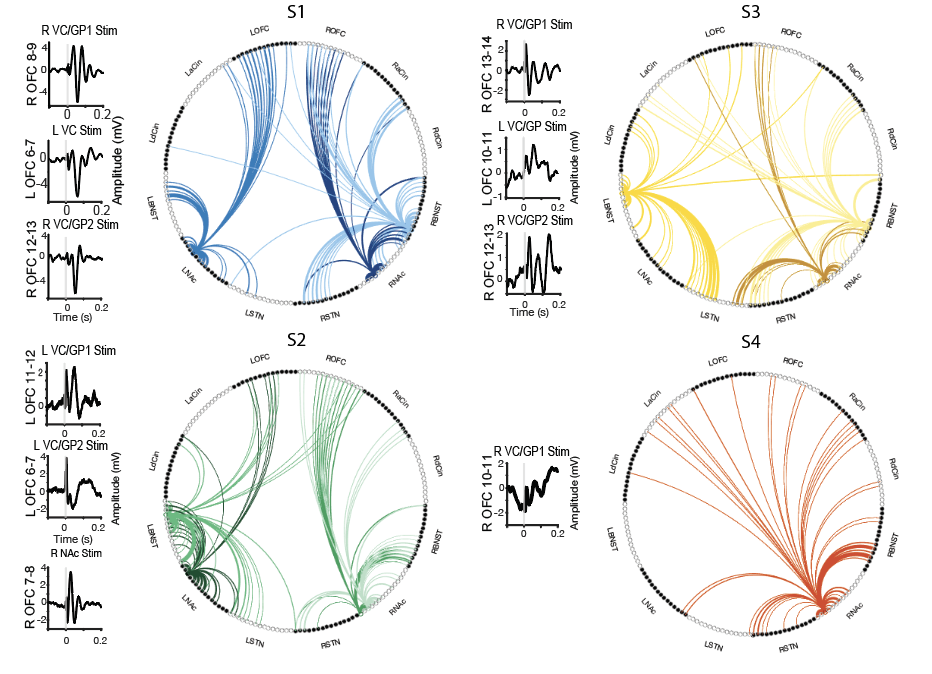

**Supp Fig 6: Cortical Evoked Potentials from Acutely Therapeutic VC/GP stimulation sites that were implanted for chronic DBS**

Example evoked potential waveforms and corresponding connectivity diagrams for acutely therapeutic VC/GP stimulation sites in subjects 1-4. Chord diagrams display cortical evoked potential magnitudes exceeding threshold (z > 1) following single-pulse stimulation of therapeutic contact pairs. Colors correspond to individual subjects: subject 1 (blue), subject 2 (green), subject 3 (yellow), and subject 4 (red).

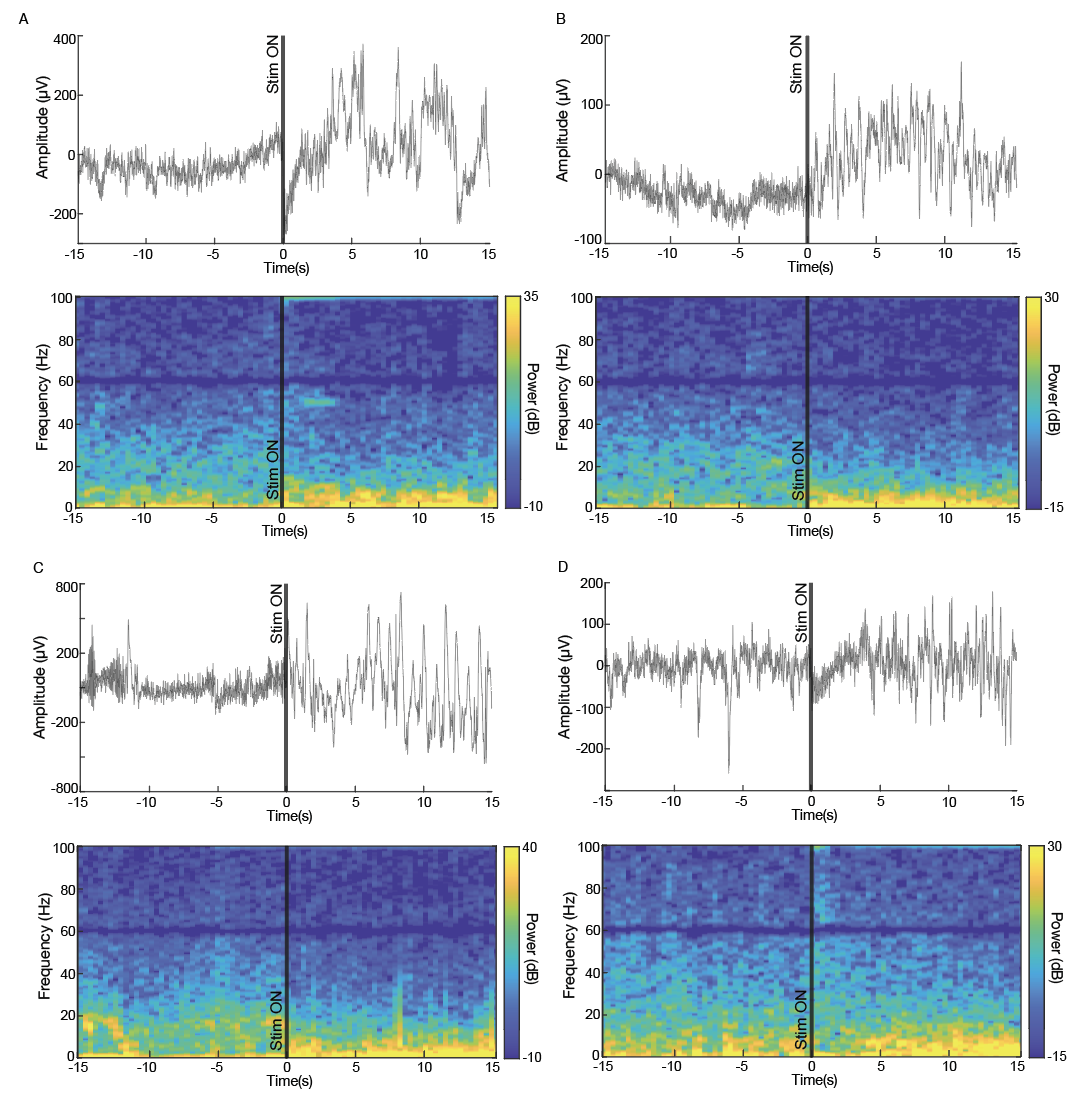

**Supp Fig 7: Example of OFC LFP Focal Slowing with Acutely Therapeutic VC Stimulation**. LFP trace and associated spectrogram of stimulation response in a representative trial in the OFC with stimulation in A) L VC in S1, B) L VC/GP in S2, C) R VC/GP2 in S3, and D) R VC/GP1 in S4.

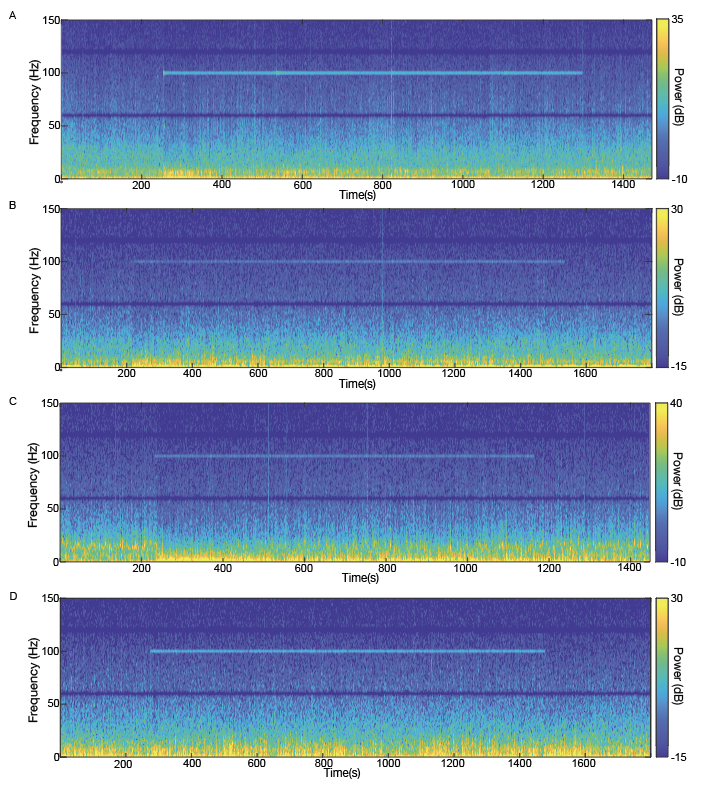

**Supp Fig 8: Examples of Prolonged LFP ‘Slowing’ After Stimulation**

Spectrogram of stimulation response i in a representative trial in the OFC with stimulation in A) L VC in S1, B) L VC/GP in S2, C) R VC/GP2 in S3, and D) R VC/GP1 in S4.

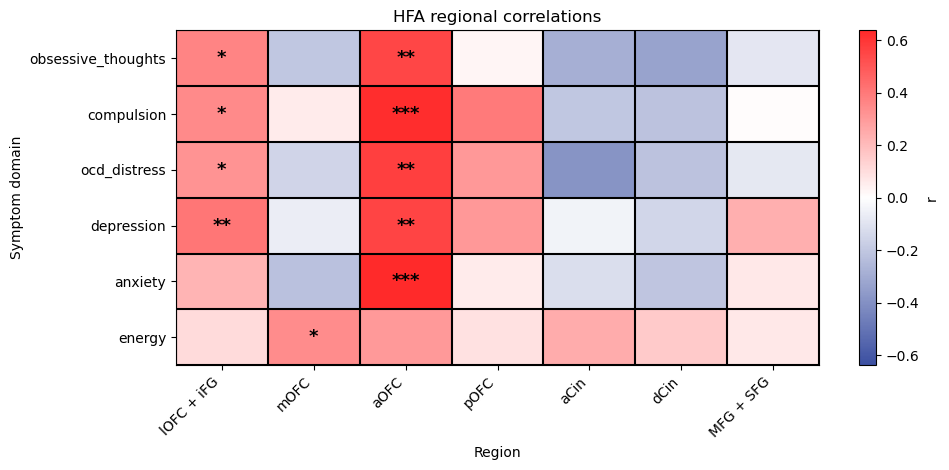

**Supp Fig 9: Correlation between Change in HFA and VAS Symptom Domains with Stimulation**

Heatmap showing correlations between changes in high-frequency activity (HFA) and VAS symptom subdomains (obsessive thoughts, compulsions, OCD distress, depression, anxiety, and energy) across stimulation sessions in subjects 1-4. HFA was quantified by comparing average power during the 30 seconds pre-stimulation to the first 5 minutes following stimulation onset and was calculated for each cortical region (lOFC+iFG, mOFC, aOFC, pOFC, aCin, dCin, and MFG+SFG). Analyses pooled both 5-minute and 20-minute stimulation trials. Asterisks denote statistical significance (*p < 0.05, **p < 0.01, ***p < 0.001).

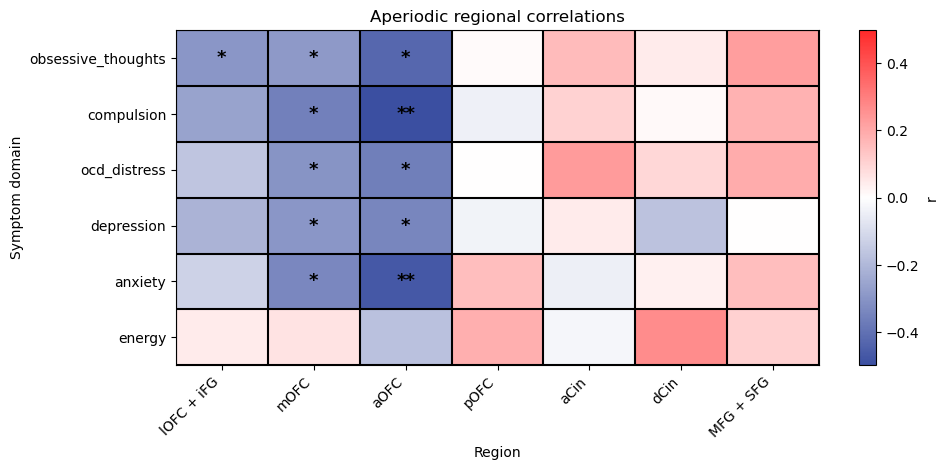

**Supp Fig 10: Correlation between Change in Aperiodic Exponent and VAS Symptom Domains with Stimulation**

Heatmap showing correlations between changes in aperiodic exponent and VAS symptom subdomains (obsessive thoughts, compulsions, OCD distress, depression, anxiety, and energy) across stimulation sessions in subjects 1-4. HFA was quantified by comparing average power during the 30 seconds pre-stimulation to the first 5 minutes following stimulation onset and was calculated for each cortical region (lOFC+iFG, mOFC, aOFC, pOFC, aCin, dCin, and MFG+SFG). Analyses pooled both 5-minute and 20-minute stimulation trials. Asterisks denote statistical significance (*p < 0.05, **p < 0.01, ***p < 0.001).

DP01

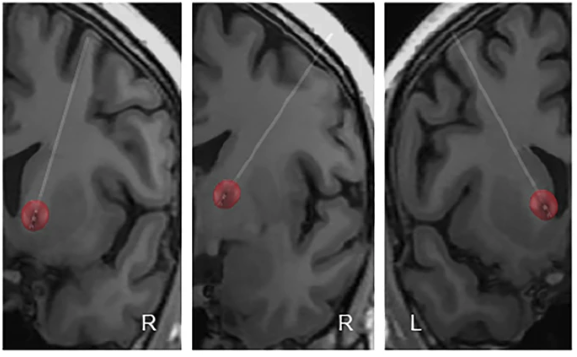

DP02

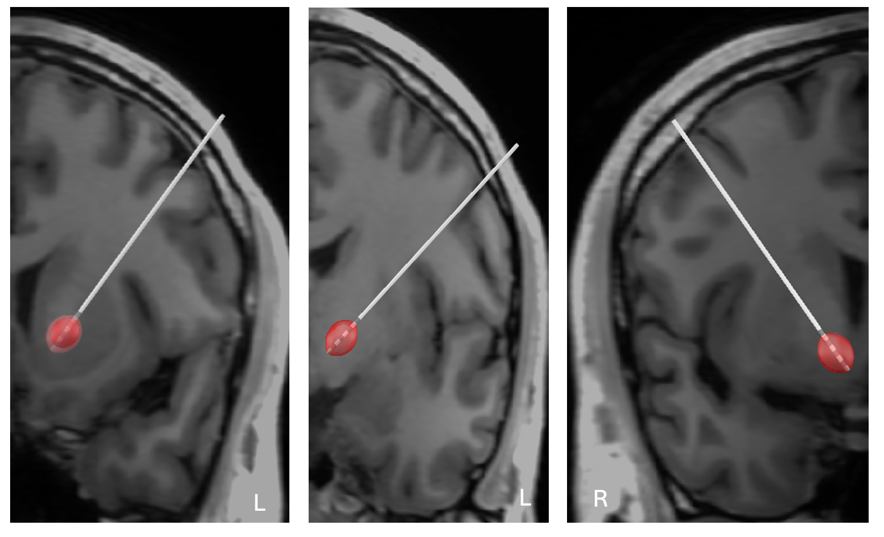

DP03

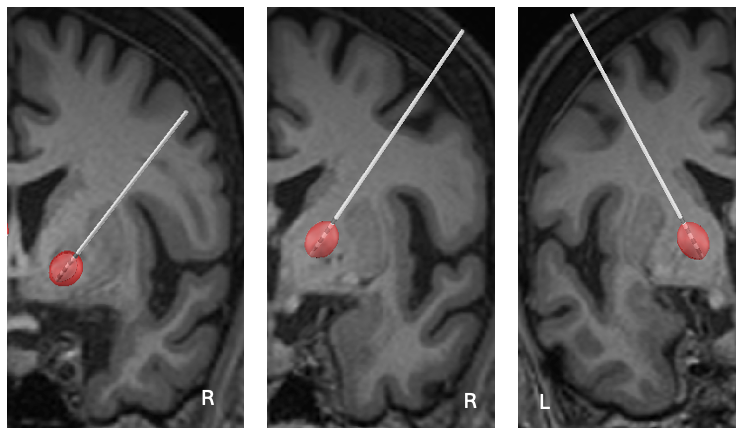

**Supp Fig 11: DBS Lead Localization Across Subjects.** DBS lead localization of permanently implanted stimulation targets in subjects 1-3 visualized using Lead-DBS.
